# Pulmonary Hypertension and the Risk of 30-day Postoperative Pulmonary Complications after Gastrointestinal Surgical or Endoscopic Procedures: A Retrospective Propensity Score Weighted Cohort Analysis

**DOI:** 10.1101/2024.02.20.24303112

**Authors:** Yoshio Tatsuoka, Zyad J. Carr, Sachidhanand Jayakumar, Hung-Mo Lin, Zili He, Adham Farroukh, Paul Heerdt

## Abstract

**Background:** Pulmonary hypertension (PH) patients are at higher risk of postoperative complications. We analyzed the association of PH with 30-day postoperative pulmonary complications (PPC).

**Methods:** A single-center propensity score overlap weighting (OW) retrospective cohort study of 164 patients with mean pulmonary artery pressure (mPAP) of >20 mmHg within 24 months of procedure and a control cohort (N=1981), undergoing elective inpatient abdominal surgery or endoscopic procedures under general anesthesia. The primary outcome was PPC, and the secondary outcomes were PPC sub-composites; respiratory failure (RF), pneumonia (PNA), aspiration pneumonia/pneumonitis (ASP), pulmonary embolism (PE), length of stay (LOS), and 30-day mortality.

**Results:** PPC was higher in the PH cohort (29.9% vs. 11.2%, p<0.001). When sub-composites were analyzed, higher rates of RF (19.3% vs. 6.6%, p<0.001) and PNA (11.2% vs. 5.7%, p=0.01) were observed. After OW, PH was still associated with higher PPC [RR 1.66, 95% CI (1.05 – 2.71), p=0.036] and increased LOS (median 8.0 days vs. 4.9 days) but not 30-day mortality. Sub-cohort analysis showed no difference in PPC between pre- and post-capillary PH patients.

**Conclusions:** After covariate balancing, PH was associated with a higher risk for PPC and prolonged LOS. This elevated PPC risk should be considered during preoperative risk assessment.

## 1. Introduction

Pulmonary hypertension (PH) is an adverse cardiopulmonary condition that is precipitated by a range of pathologic insults affecting the blood vessels of the lung [1]. Currently, a definitive diagnosis of PH requires right heart catheterization and a measured mean pulmonary arterial pressure (mPAP) of > 20 mmHg with hemodynamic etiology (pre-capillary, post-capillary, or combined pre- and post-capillary) defined by measurement of pulmonary arterial occlusion pressure (PAOP) and calculation of pulmonary vascular resistance (PVR). Adverse perioperative outcomes have been observed in patients with PH, but it is less clear whether there is an association with increased 30-day postoperative pulmonary complications (PPC) which occur with an incidence of 1 to 23% in surgical populations [2,3]. In 2005, Ramakrishna and colleagues observed that of 144 PH patients who survived surgery, 60 (42%) had one or more perioperative morbid events, of which respiratory failure (28%), cardiac arrhythmia (12%) and congestive heart failure (11%) were most common. Patients undergoing intermediate to high-risk surgery were observed to have a 2.5-fold increase in morbid events when compared to low-risk procedures. Increased post-anesthesia care unit respiratory depressive episodes and increased in-hospital PPC in PH patients have been observed after noncardiac surgery [4]. Similarly, an increased risk for postoperative in-hospital pneumonia was observed in PH patients after cardiac valve surgery [5]. Given the trend toward shorter length of stay, it is important to characterize predictive factors for PPC in a 30-day timeframe to better identify perioperative risk [6]. In this study, we sought to estimate and compare 30-day PPC in a population of PH (mPMP >20 mmHg) and controls undergoing abdominal surgical and endoscopic procedures. We selected abdominal procedures to avoid the higher 30-day PPC incidence associated with thoracic and cardiac procedures. We hypothesized that, even after covariate balancing, the diagnosis of PH would be associated with increased risk of 30-day PPC, as demonstrated by the Agency for Healthcare Research and Quality (AHRQ) PPC composite outcome. For exploratory secondary outcome analyses, we investigated its association with categorized sub-composites of infectious pneumonia (PNA), aspiration pneumonia/pneumonitis (ASP), respiratory failure (RF), pulmonary embolism (PE), and 30-day mortality. Furthermore, we compared pre-capillary [Pulmonary Artery Occlusion Pressure (PAOP) <15 mmHg] and post-capillary (PAOP >15 mmHg) PH to explore differences between these distinct PH etiologies.

## 2. Materials and Methods

This retrospective cohort analysis was approved by the Yale University Institutional Review Board (IRB#2000032516) and received a waiver of informed consent. We strove to adhere to the Strengthening the Reporting of Observational Studies in Epidemiology (STROBE) Statement on reporting observational studies.

### Study Population

Using a retrospective single-center comparative cohort design, we identified adult patients (18-90 years old) with International Classification of Disease version 10 (ICD-10) codes for PH (I27.0, I27.2) and American Society of Anesthesiologists Physical Status (ASA-PS) I to IV undergoing elective inpatient upper and lower abdominal surgery or gastrointestinal endoscopic procedures at a large quaternary care facility located in the United States between Jan 1, 2013, and Jan 1, 2020. The selection of these dates coincides with the introduction of a modern electronic record at the institution (Epic systems, Verona WI, USA). Performed procedures were under general anesthesia with or without endotracheal intubation. After the initial identification of 2,792 PH patients, we performed a manual electronic chart review and excluded patients without right heart catheterization data consistent with PH (mPAP >20 mmHg) within 24 months of procedure. 167 patients were identified in the PH cohort. A control cohort of 2005 patients with similar ages, biological sex, and procedures were identified for comparison purposes. Due to incomplete outcomes, 27 (24; control; 3; PH) patients were removed from the final propensity score weighting analysis, and a total of 164 patients in the PH cohort and 1981 patients in the control cohort were identified (Figure 1).

**Figure 1.**
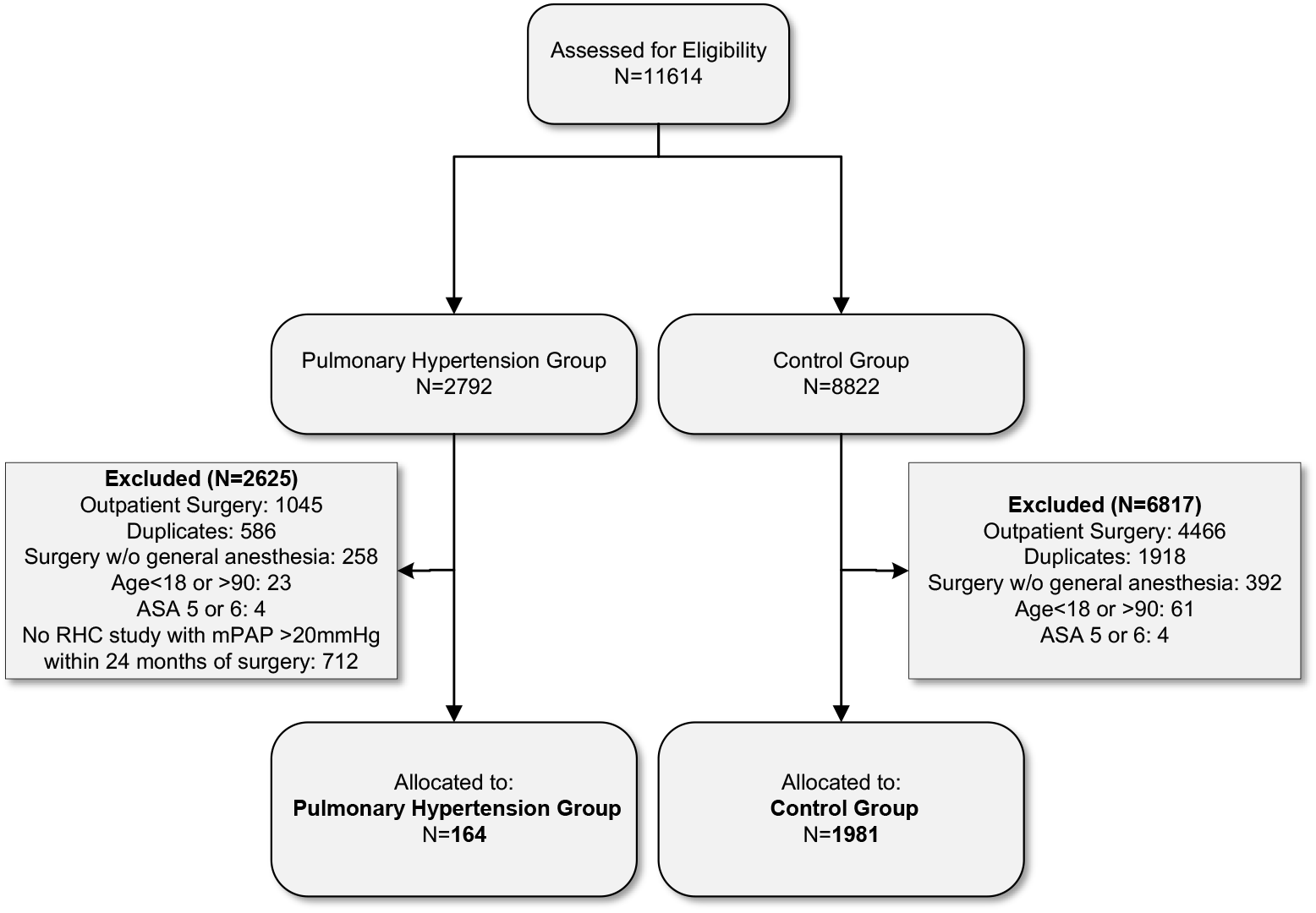
A flow diagram of patient selection criteria. Abbreviations: ASA: American Society of Anesthesiologists Physical Status; RHC: right heart catheterization; mPAP: mean pulmonary artery pressure.

### Study Data Abstraction

We extracted demographical data, including age, biological sex, race, body mass index (BMI), Elixhauser comorbidity variables [7], and hospital site. We defined PH as a mean pulmonary arterial pressure (mPAP) >20 mmHg on right heart catheterization (RHC) within two years of surgical procedure. For the purposes of procedural risk matching, procedure severity was defined using the Surgical Outcome Risk Tool (SORT) v2 [8]. We used the number of Elixhauser comorbidity variables as a measure of frailty [9,10]. 30-day PPC was defined by coding guidance provided by the AHRQ PPC composite outcome (Appendix Supplemental Table). We further divided the constituent codes into four separate sub-composites for detailed analysis of respiratory failure (RF), infectious pneumonia (PNA), aspiration pneumonia/pneumonitis (ASP), and pulmonary embolism (PE) to better understand their individual contribution to PPC in the study population. 30-day mortality, length of stay (LOS), and mortality-free discharge were also analyzed for both cohorts. Given the distinct differences between PH subtypes, we further divided the PH cohort into pre-capillary PH (PAOP <15 mmHg) and post-capillary PH (PAOP > 15 mmHg) to analyze the primary outcome.

### Outcomes

Our primary outcome was PPC, and secondary outcomes of RF, PNA, ASP, PE, LOS, and 30-day mortality. In a sub cohort analysis, we compared pre-capillary PH and post-capillary PH to the primary and secondary outcomes.

### Statistical Analysis

After verification of study variables, descriptive statistics were reported as mean and standard deviation (SD) or median and interquartile range [IQR] for continuous variables and frequency (%) for categorical variables. Initial assessment of the PH and control cohort found significant systematic differences in baseline characteristics. We utilized the overlap weighting (OW) approach to optimize precise balance of the baseline covariates. OW is a propensity score (PS) method that attempts to mimic important attributes of randomized clinical trials. Briefly, OW assigns a weight to each patient that is proportional to the probability of that patient belonging to the opposite group. We chose the OW method because of substantial baseline differences between PH and control cohorts. OW has desirable statistical properties and leads to an exact balance on the mean of every measured covariate when the PS is estimated by a logistic regression [11]. The target population is the group of patients for whom the conclusions are drawn [12]; in our case, patients who undergo elective abdominal surgery or endoscopic procedures under general anesthesia. Due to incomplete outcomes, 27 (24; control; 3; PH) patients were removed before the final OW analysis. We fit a logistic regression model to estimate the PH propensity scores using baseline covariates - age, biological sex, race, BMI, number of Elixhauser comorbidities, procedural severity, and hospital location. The OW were then generated and incorporated into all subsequent outcome analyses. Finally, we evaluated the group differences based on the standardized mean differences (SMD), where an SMD<0.1 is usually deemed successful in achieving covariate balancing.

In the outcome analysis, for the binary outcomes PPC, RF, PNA, ASP, PE, and 30-day mortality, we used weighted log binomial regression to obtain the risk ratio (RR) for RHC group compared to control. For the LOS outcome, we used weighted survival analysis, in which mortality-free discharge was the event and in-hospital death patients were censored. Weighted Cox proportional hazards model was used to obtain the hazard ratio (HR) of the mortality-free discharge for the PH cohort. Under this setting, a HR<1 indicates a lower chance of mortality-free discharge. For the sub-cohort, no p-value adjustment was made due to the small sample size and lack of significant findings. We accepted a p-value of <0.05 to reject the null hypothesis. Data management and statistical analyses was carried out using R version 4.3.0.

## 3. Results

### 3.1.1. Baseline characteristics of PH and control cohorts

The baseline characteristics are described in Table 1. The study included the PH cohort (N=167) and the control cohort (N=2005). The PH cohort was older (62 vs. 57 years old, p<0.001), had more non-white ethnicity (44.5 vs. 26.8%, p<0.001), higher number of Elixhauser comorbidities (6.9 vs. 3.2, p<0.001), and reduced procedural severity (Minor or intermediate 68.3 vs. 32.5%; Major 15.6 vs. 43.0%; Xmajor/complex 16.5% vs. 24.5%; p<0.001). For PH patients, more procedures were performed at the large quaternary care facility (72.6% vs. 61.9%, p= 0.02). Prior to propensity score overlap weighting adjustment, the PH cohort had higher 30-day PPC incidence (29.9% vs. 11.2%, p<0.001). Regarding unadjusted secondary outcomes, PH cohort was associated with significantly higher rates of RF (19.3% vs. 6.6%, p<0.001), and PNA (11.2% vs. 5.7%, p=0.010), but not ASP (3.7% vs. 1.7%, p=0.118), PE (3.1% vs. 1.5%, p=0.181) nor 30-day mortality (3.0% vs. 2.5%, p=0.604).

**Table 1.**
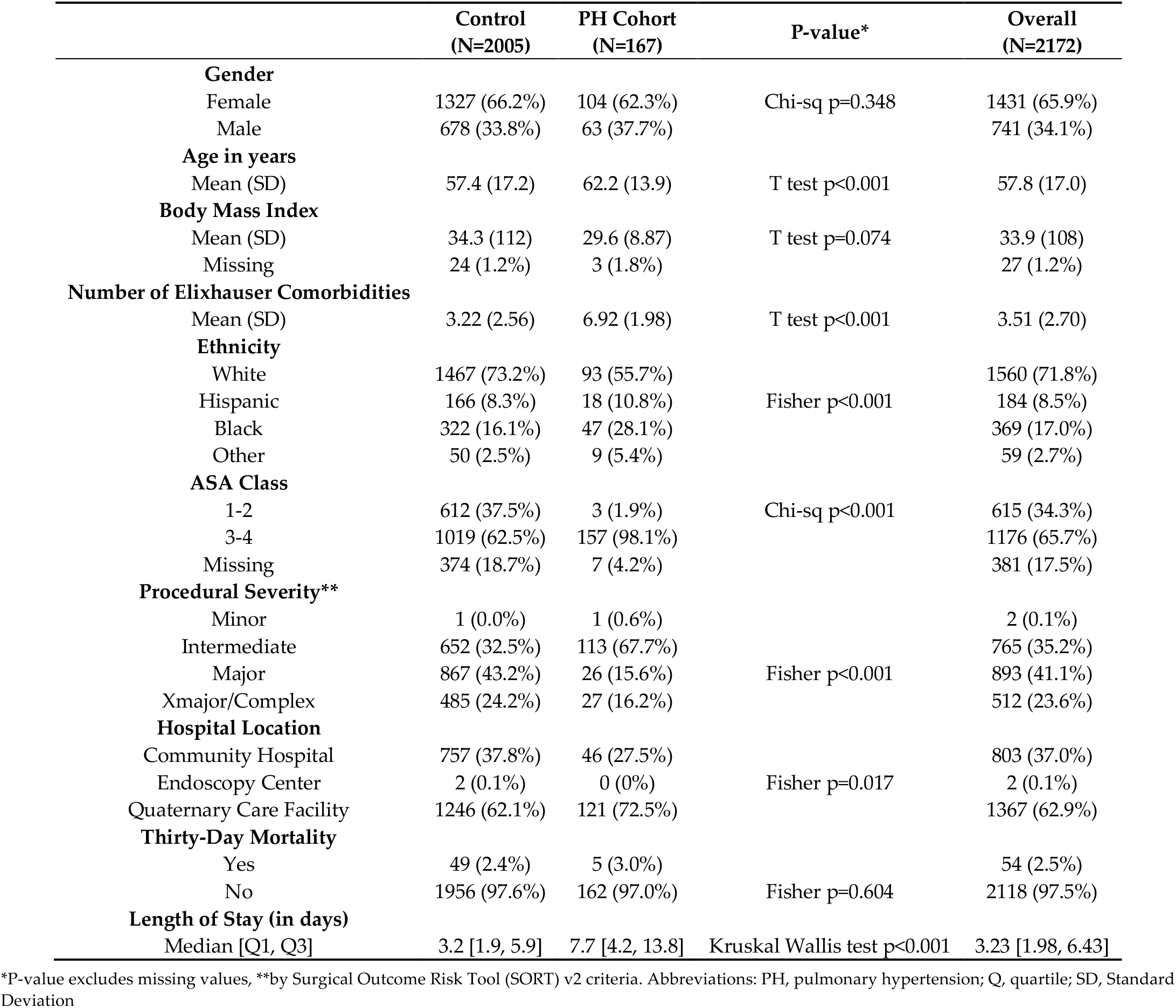
Comparisons of the Baseline Characteristics between Control vs. PH Cohort.

|  | Control<br>(N=2005) | PH Cohort<br>(N=167) | P-value* | Overall<br>(N=2172) |
| --- | --- | --- | --- | --- |
| **Gender** |  |  |  |  |
| Female | 1327 (66.2%) | 104 (62.3%) | Chi-sq p=0.348 | 1431 (65.9%) |
| Male | 678 (33.8%) | 63 (37.7%) |  | 741 (34.1%) |
| **Age in years** |  |  |  |  |
| Mean (SD) | 57.4 (17.2) | 62.2 (13.9) | T test p<0.001 | 57.8 (17.0) |
| **Body Mass Index** |  |  |  |  |
| Mean (SD) | 34.3 (112) | 29.6 (8.87) | T test p=0.074 | 33.9 (108) |
| Missing | 24 (1.2%) | 3 (1.8%) |  | 27 (1.2%) |
| **Number of Elixhauser Comorbidities** |  |  |  |  |
| Mean (SD) | 3.22 (2.56) | 6.92 (1.98) | T test p<0.001 | 3.51 (2.70) |
| **Ethnicity** |  |  |  |  |
| White | 1467 (73.2%) | 93 (55.7%) | Fisher p<0.001 | 1560 (71.8%) |
| Hispanic | 166 (8.3%) | 18 (10.8%) |  | 184 (8.5%) |
| Black | 322 (16.1%) | 47 (28.1%) |  | 369 (17.0%) |
| Other | 50 (2.5%) | 9 (5.4%) |  | 59 (2.7%) |
| **ASA Class** |  |  |  |  |
| 1-2 | 612 (37.5%) | 3 (1.9%) | Chi-sq p<0.001 | 615 (34.3%) |
| 3-4 | 1019 (62.5%) | 157 (98.1%) |  | 1176 (65.7%) |
| Missing | 374 (18.7%) | 7 (4.2%) |  | 381 (17.5%) |
| **Procedural Severity\*\*** |  |  |  |  |
| Minor | 1 (0.0%) | 1 (0.6%) | Fisher p<0.001 | 2 (0.1%) |
| Intermediate | 652 (32.5%) | 113 (67.7%) |  | 765 (35.2%) |
| Major | 867 (43.2%) | 26 (15.6%) |  | 893 (41.1%) |
| Xmajor/Complex | 485 (24.2%) | 27 (16.2%) |  | 512 (23.6%) |
| **Hospital Location** |  |  |  |  |
| Community Hospital | 757 (37.8%) | 46 (27.5%) | Fisher p=0.017 | 803 (37.0%) |
| Endoscopy Center | 2 (0.1%) | 0 (0%) |  | 2 (0.1%) |
| Quaternary Care Facility | 1246 (62.1%) | 121 (72.5%) |  | 1367 (62.9%) |
| **Thirty-Day Mortality** |  |  |  |  |
| Yes | 49 (2.4%) | 5 (3.0%) | Fisher p=0.604 | 54 (2.5%) |
| No | 1956 (97.6%) | 162 (97.0%) |  | 2118 (97.5%) |
| **Length of Stay (in days)** |  |  |  |  |
| Median [Q1, Q3] | 3.2 [1.9, 5.9] | 7.7 [4.2, 13.8] | Kruskal Wallis test p<0.001 | 3.23 [1.98, 6.43] |

\*P-value excludes missing values, \*\*by Surgical Outcome Risk Tool (SORT) v2 criteria. Abbreviations: PH, pulmonary hypertension; Q, quartile; SD, Standard Deviation

### 3.1.2. Overlap Weighted Outcome Analysis

Comparison between the PH cohort and the control cohort before and after OW adjustment is described in Table 2. 27 (24; control; 3; PH) patients were removed due to missing variables for OW analysis. Post-adjustment, the SMD for each characteristic is all <0.01, indicating successful covariate balancing. Table 3 shows the findings after propensity weighted matching. PH was associated with increased risk for 30-day PPC [Risk Ratio (RR) 1.66, 95% CI (1.05 – 2.71), p=0.036]. There were no differences in sub-composite RF [RR 1.68, 95% CI (0.90 – 3.28), p=0.109], PNA [RR 1.21, 95% CI (0.57 – 2.65), p=0.615], ASP [RR 1.63, 95% CI (0.38 – 8.28), p=0.514], and PE [RR 1.20, 95% CI (0.29 – 5.28), p=0.794]. Survival analysis demonstrates that PH was associated with increased LOS [in days, median 8.0 vs. 4.9, 95% CI (6.8 – 9.1) vs. 4.9 (4.4 – 5.2)]; mortality-free discharge: [HR 0.63, 95% CI (0.53 – 0.77), p=<0.001]. These conclusions remain the same even after adjusting the p-values for multiple secondary outcomes.

**Table 2.** Comparison of PH versus Control before and after Propensity Score Overlap Weighting Adjustment.

| Variables | Before Overlap Weighting |  |  | After Overlap Weighting |  |  |
| --- | --- | --- | --- | --- | --- | --- |
|  | Control<br>(N=1981)* | PH<br>(N=164)* | SMD | Control<br>(N=1981)* | PH<br>(N=164)* | SMD |
| **Age (years), mean (SD)** | 57.4 (17.18) | 62.02 (13.72) | 0.29 | 62.62 (15.65) | 62.62 (14.10) | **< 0.01** |
| **Gender** |  |  | 0.08 |  |  | **< 0.01** |
| Female | 1310 (66.1) | 102.0 (62.2) |  | 72.4 (60.8) | 72.4 (60.8) |  |
| Male | 671.0 (33.9) | 62.0 (37.8) |  | 46.6 (39.2) | 46.6 (39.2) |  |
| **Race** |  |  | 0.39 |  |  | **< 0.01** |
| White, Non-Hispanic | 1450 (73.2) | 91.0 (55.5) |  | 71.6 (60.1) | 71.6 (60.1) |  |
| Hispanic | 165 (8.3) | 18 (11.0) |  | 12.0 (10.1) | 12.0 (10.1) |  |
| Black | 317 (16.0) | 46.0 (28.0) |  | 30.1 (25.3) | 30.1 (25.3) |  |
| Other | 49.0 (2.5) | 9.0 (5.5) |  | 5.3 (4.5) | 5.3 (4.5) |  |
| **BMI, mean (SD)** | 34.2 (112.4) | 29.6 (8.8) | 0.06 | 29.6 (15.1) | 29.6 (9.0) | **< 0.01** |
| **Number of Elixhauser comorbidities, mean (SD)** | 3.2 (2.55) | 6.9 (1.97) | 1.64 | 6.5 (2.77) | 6.5 (1.83) | **< 0.01** |
| **Hospital Location** |  |  | 0.23 |  |  | **< 0.01** |
| Community Hospital | 753 (38.0) | 45 (27.4) |  | 36.3 (30.5) | 36.3 (30.5) |  |
| Endoscopy Center | 2.0 (0.1) | 0.0 (0.0) |  | 0.0 (0.0) | 0.0 (0.0) |  |
| Quaternary Care Facility | 1226.0 (61.9) | 119 (72.6) |  | 82.7 (69.5) | 82.7 (69.5) |  |
| **Procedure Severity** |  |  | 0.80 |  |  | **< 0.01** |
| Minor or Intermediate | 644 (32.5) | 112 (68.3) |  | 76 (63.9) | 76 (63.9) |  |
| Major | 852 (43.0) | 25 (15.2) |  | 21 (18.0) | 21 (18.0) |  |
| Xmajor/Complex | 485 (24.5) | 27 (16.5) |  | 21 (18.2) | 21 (18.2) |  |

Values are N (%) unless otherwise indicated. \*27 (24 in control cohort, 3 PH cohort) patients were removed due to missing variables for propensity score overlap weighting analysis. Abbreviations: PH, pulmonary hypertension; SMD; standardized mean difference; BMI, body mass index (kg/m<sup>2</sup>)

**Table 3.** Overlap Weighed Propensity Model Findings of Primary and Secondary Outcomes in the PH cohort.

| Outcome | Unadjusted* |  |  | Propensity Overlap Weighted Model** |  |  |
| --- | --- | --- | --- | --- | --- | --- |
|  | PH | Control | P-value | RR | 95% CI | P-value |
| **PPC** | 49 (29.9%) | 221 (11.2%) | **<0.001** | 1.66 | 1.05 – 2.71 | **0.036** |
| **RF** | 31(19.3%) | 131 (6.6%) | **<0.001** | 1.68 | 0.90 – 3.28 | 0.109 |
| **PNA** | 18 (11.2%) | 113 (5.7%) | **0.010** | 1.21 | 0.57 – 2.65 | 0.615 |
| **ASP** | 6 (3.7%) | 34 (1.7%) | 0.118 (f) | 1.63 | 0.38 – 8.28 | 0.514 |
| **PE** | 5 (3.1%) | 30 (1.5%) | 0.181 (f) | 1.2 | 0.29 – 5.28 | 0.794 |
| **30-day Mortality** | 5 (3.0%) | 49 (2.5%) | 0.604 (f) | 1.23 | 0.43 - 2.76 | 0.651 |
| **Length of stay, median days (IQR)** | 8 (6.8, 9.1) | 4.9 (4.4, 5.2) | **<0.001** | 0.63 | 0.53 - 0.77 | **<0.001** |

\*Chi-Square test; p-value excludes missing values, \*\*Propensity weighted by the following variables: age, biological sex, race, BMI, procedural severity, and number of Elixhauser covariates. (f) Fisher's Exact test applied. Abbreviations: PH, pulmonary hypertension; PPC, postoperative pulmonary complications; RF, respiratory failure; PNA, infectious pneumonia; ASP, aspiration pneumonia/pneumonitis; PE, pulmonary embolism; RR, relative risk; CI, confidence interval; IQR, interquartile range; BMI, body mass index.

### 3.1.3. Sub-cohort Analysis: Comparisons of pre-capillary vs. post-capillary PH

Baseline characteristics of the sub-cohorts and OW model outcomes are described in Table 4. To analyze for between-group differences, the PH cohort was divided into pre-capillary PH (N=47; mPAP >20 mmHg and PAOP <15 mmHg) and post-capillary PH (N=116, mPAP >20 mmHg and PAOP >15 mmHg). We observed no difference in the primary outcome of PPC (31.9% vs. 29.3%, p=0.89) nor secondary outcomes of RF (21.3% vs. 18.6%, p=0.86), PNA (4.3% vs. 14.2%, p=0.126), ASP (6.4% vs. 2.7%, p=0.36), and PE (2.1% vs. 3.5%, p=1.0).

**Table 4.** Baseline Characteristics and Postoperative Outcomes of the PH Cohort and Comparison of Outcome between Pre- and Post-capillary PH Groups.

| Variable | PH group | Pre-capillary | Post-capillary | P-value* |
| --- | --- | --- | --- | --- |
| Age (years) | 63.2 (13.9) | 59.88 (14.8) | 63.0 (13.5) | 0.18 |
| BMI (kg/m<sup>2</sup>) | 29.6 (8.9) | 27.8 (10.4) | 30.2 (8.0) | 0.12 |
| **Echocardiography** |  |  |  |  |
| Right Ventricular Systolic Pressure (mmHg) | 52.9 (20.6) | 54.3 (20.3) | 52.2 (20.9) | 0.59 |
| **Right Heart Catheterization** |  |  |  |  |
| Mean Right Atrial Pressure (mmHg) | 13.2 (7.7) | 9.5 (5.9) | 14.6 (8.0) | <0.001 |
| Right Ventricular Systolic Pressure (mmHg) | 53.8 (18.9) | 51.9 (19.8) | 54.5 (18.6) | 0.42 |
| Pulmonary Artery Systolic Pressure (mmHg) | 53.3 (18.4) | 51.2 (19.9) | 54.0 (17.7) | 0.37 |
| Pulmonary Artery Diastolic Pressure (mmHg) | 25.0 (9.1) | 21.5 (8.6) | 26.4 (8.9) | 0.002 |
| Pulmonary Artery Mean Pressure (mmHg) | 35.7 (11.6) | 33.2 (11.4) | 36.7 (11.4) | 0.08 |
| Pulmonary Artery Occlusion Pressure (mmHg) | 19.1 (7.3) | 11.35 (2.5) | 22.2 (6.2) | <0.001 |
| Pulmonary Artery Oxygen Saturation (%) | 64.3 (9.6) | 65.5 (8.5) | 64.0 (9.8) | 0.39 |
| Fick Cardiac Output (L/min) | 5.9 (2.5) | 5.2 (1.9) | 6.2 (2.6) | 0.02 |
| Calculated Cardiac Index | 3.1 (1.2) | 3.0 (1.2) | 3.1 (1.1) | 0.60 |
| **Primary and Secondary Outcomes\*\*** |  | **Pre-capillary**<br>**(N=47)** | **Post-capillary**<br>**(N=116)** | **P-value** |
| PPC |  | 15 (31.9%) | 34 (29.3%) | 0.889 |
| RF |  | 10 (21.3%) | 21 (18.6%) | 0.863 |
| PNA |  | 2 (4.3%) | 16 (14.2%) | 0.126 |
| ASP |  | 3 (6.4%) | 3 (2.7%) | 0.36 |
| PE |  | 1 (2.1%) | 4 (3.5%) | 1 |

Figures in baseline characteristics sections are mean, followed by SD in parentheses unless otherwise indicated. \*2-side P-value after unadjusted t-test, equal variances assumed \*\*4 patients were excluded for missing pulmonary artery occlusion pressure. Abbreviations: PH, pulmonary hypertension; PPC, postoperative pulmonary complications; RF, respiratory failure; PNA, infectious pneumonia; ASP, aspiration pneumonia/pneumonitis; PE, pulmonary embolism, SD, standard deviation.

## 4. Discussion

In our study, we identified an unadjusted 29.9% risk for PPC in the PH cohort vs. 11.2% in the control cohort after inpatient abdominal surgical and endoscopic procedures under general anesthesia. After covariate balancing, PH was associated with a 66% increase in the risk for PPC in the 30 days following procedure. Secondly, this increase in 30-day PPC increased LOS but not 30-day mortality. It has been estimated that 1% of the global population and up to 10% of those older than 65 years of age have PH, making it imperative to identify perioperative outcomes associated with this pathological condition [13]. Using a detailed composite measure of PPC, secondary sub-composite analysis did not provide any guidance toward specific categories that most strongly contributed to the development of PPC in PH patients. PH patients demonstrated higher incidence across all sub-composites when compared to controls (Figure 2). In addition to the short-term risks of perioperative administration of respiratory depressants and surgery-related compromise of respiratory mechanics, this lack of discrimination suggests two PH-related physiological consequences may contribute to a generalized 30-day PPC risk. PH patients demonstrate an underlying dysregulated immune response [14]. Dysregulated inflammation has been associated with an increased propensity for perioperative complications, multiple organ dysfunction, and increased postoperative complications [15,16]. In a study of patients with pulmonary arterial hypertension and chronic thromboembolic-derived PH, circulating c-reactive protein (CRP), a marker for generalized inflammation, is significantly higher in PH compared to controls [17]. Furthermore, it is understood that underlying dysregulated immune responses may increase susceptibility to infectious pneumonia and favor the development of thromboembolic phenomenon [18]. This may be consistent with our observation of a 30-day postoperative pneumonia and pulmonary embolism prevalence nearly twice that of controls. Secondly, it may point to a general susceptibility to deleterious fluid shifts in the perioperative period. It is well understood that right ventricular dysfunction is common in PH patients during physiological stress [19,20]. Increasing serum natriuretic peptide, a validated measure of HF severity, has been associated with worse weaning outcomes in postoperative patients, is a measure of risk stratification in pulmonary arterial hypertension, and a biomarker for PH-related right ventricular dysfunction [21-24]. Perioperative volume overload and positive cumulative fluid balance have been associated with increased morbidity and mortality risk in both general and surgical subpopulations [25]. Although limited by the small sample size, we did not observe a significant increase in 30-day mortality. 30-day mortality after elective non-cardiac surgery in PH patients has been reported between 2% and 18% [2,26,27]. Both short- and long-term postoperative mortality has been observed to increase in patients who develop PPC [28], and increased 90-day mortality risk has been demonstrated after abdominal surgery [29]. However, LOS was nearly 2-fold that of controls in the PH cohort, suggesting that although failure to rescue from perioperative complications is low, it does increase healthcare utilization. In the survival analysis where mortality-free hospital discharge was compared, the PH cohort had a significantly lower likelihood of mortality-free discharge after OW adjustment. Thus, it is likely that PH increased the complexity and cost of required medical care in our study cohort. We did not observe a difference in PPC between pre-capillary and post-capillary PH. This was a limited observation, given the small sample size of our study population, but surprising, given the higher generalized morbidity and poorer survival exhibited by patients with pre-capillary PH [30,31].

**Figure 2.**
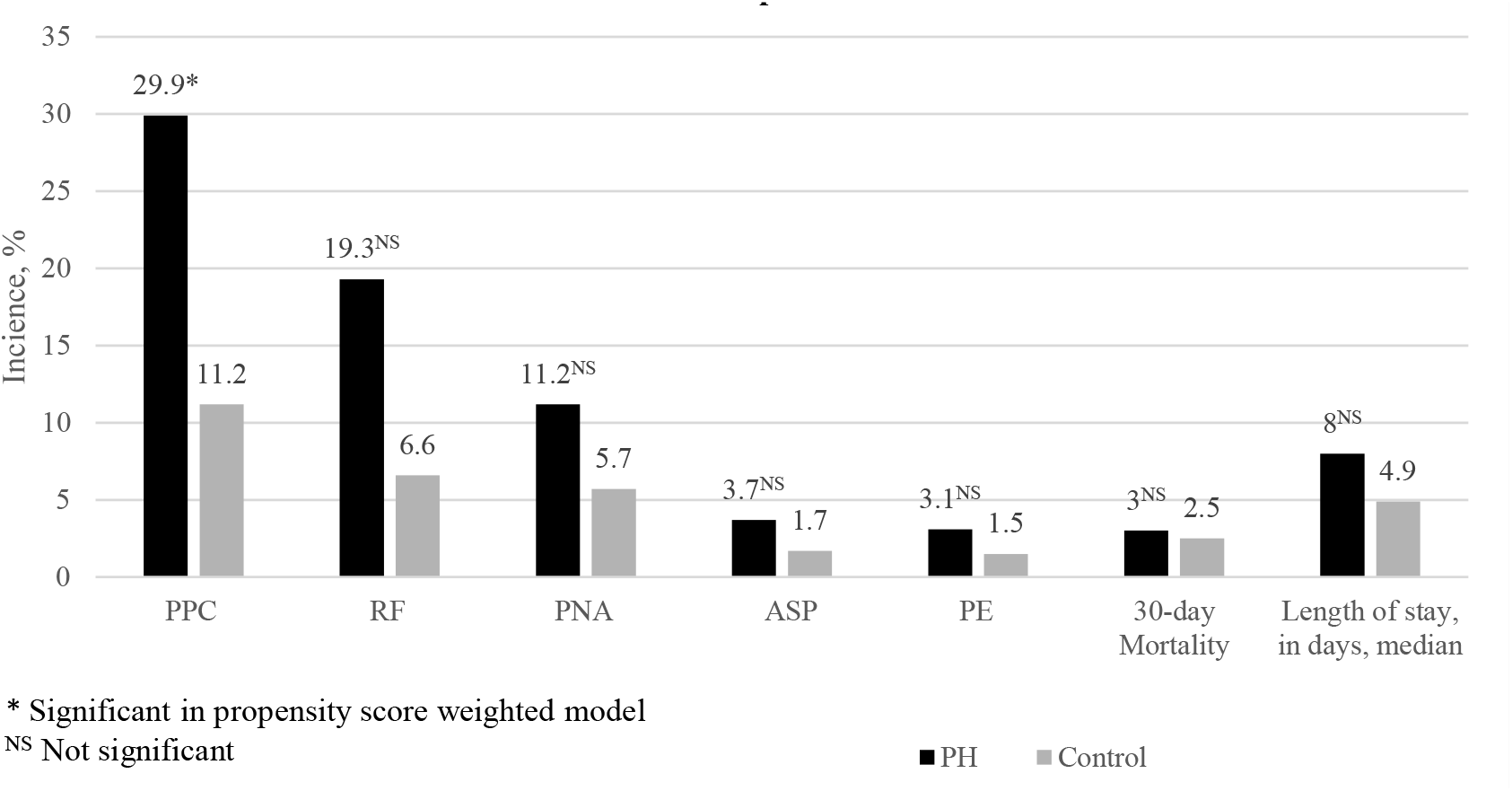
A Comparison of PH vs. Control cohorts Unadjusted Incidence of PPC and Sub-composites.

### Limitations

Our findings should be considered in the context of the potential confounding factors associated with retrospective cohort studies. To minimize convenience sampling, we performed random frequency matching stratified by age, biological sex, and procedure. We attempted to reduce misclassification bias by initial use of ICD-10 data followed by manual extraction of right heart catheterization data utilizing a cutoff value of mPAP >20 mmHg for PH cohort inclusion [32]. Furthermore, we used a 24-month window for RHC data, potentially reducing diagnostic precision for PH, although there is no available guidance regarding RHC surveillance criteria in PH populations. Due to incomplete data, we were unable to capture diagnostic group data for clinical classification of PH in our study population. Similarly, we were only able to utilize PAOP < or >15 mmHg to define pre- and post-capillary groups for exploratory sub-cohort analysis due to incomplete reporting of pulmonary vascular residence on RHC reports. Although OW can achieve highly precise balancing and minimize between-group variance, it cannot adjust for unmeasured patient characteristics or unknown confounding variables at the time of the study.

## 5. Conclusions

This study reports that PH was associated with a 66% increased risk for 30-day PPC in patients undergoing abdominal surgical and endoscopic procedures under general anesthesia. We did not observe differences between pre-capillary and post-capillary groups on the primary and secondary outcomes. The diagnosis of PH was associated with a significant increase in LOS after abdominal procedures, suggesting that postoperative care utilization is increased in this population. These observations bring attention to the extended perioperative PPC risk profile in PH, and highlight the need for identification of modifiable risk factors and further evidence-based protocolized postoperative care.

## Data Availability

All data produced in the present study are available upon reasonable request to the authors

## Author Contributions

Conceptualization and methodology, Y.T, Z.C., S.J., and P.H.; formal analysis and validation, Z.C., H.L., and Z.H.; investigation, Y.T, Z.C., and S.J.; data curation, Y.T, Z.C., S.J., and A.F.; writing—original draft preparation, Y.T, Z.C.; writing—review and editing, Y.T, Z.C., S.J., H.L., Z.H., A.F., and P.H.; visualization, Y.T, Z.C., A.F.; supervision, Z.C., H.L., and P.H.; project administration, Y.T., Z.C.; funding acquisition, none. All authors have read and agreed to the published version of the manuscript.

## Institutional Review Board Statement

The study was conducted in accordance with the Declaration of Helsinki, and approved by the Institutional Review Board of Yale University (protocol code 2000032516 and date of approval 3/19/2022).

## Informed Consent Statement

Patient consent was waived since this is a retrospective single-center study utilizing a de-identified database with minimal potential risk for patients.

## Data Availability Statement

We do not have publicly archived datasets analyzed or generated during the study.

### Appendix

#### Appendix Supplementary Table

Definition of postoperative pulmonary complications (PPC) and its sub-composites: respiratory failure (RF), infectious pneumonia (PNA), aspiration pneumonia/pneumonitis (ASP), and pulmonary embolism (PE) using International Classification of Disease version 10 (ICD-10) codes provided by the Agency for Healthcare Research and Quality (AHRQ).

| AHRQ-PPC | ICD-10 | Sub-composite Inclusion |
| --- | --- | --- |
| Transfusion-related acute lung injury | J95.84 | AHRQ-RF |
| Respiratory complications not elsewhere classified | J95.85 | AHRQ-RF |
| Ventilator associated pneumonia | J95.86 | AHRQ-PNA |
| Post-procedural aspiration pneumonia | J95.87 | AHRQ-ASP |
| Other complication of ventilator | J95.88 | AHRQ-RF |
| Other intraoperative complications of respiratory system, not elsewhere classified | J95.89 | AHRQ-RF |
| Other postprocedural complications and disorders of respiratory system, not elsewhere classified | J95.90 | AHRQ-RF |
| Acute respiratory failure, unspecified whether with hypoxia or hypercapnia | J95.91 | AHRQ-RF |
| Respiratory failure, unspecified, unspecified whether with hypoxia or hypercapnia | J95.92 | AHRQ-RF |
| Respiratory failure, unspecified with hypoxia | J95.93 | AHRQ-RF |
| Respiratory failure, unspecified with hypercapnia | J95.94 | AHRQ-RF |
| Acute and chronic respiratory failure, unspecified whether with hypoxia or hypercapnia | J95.95 | AHRQ-RF |
| Acute and chronic respiratory failure with hypoxia | J95.96 | AHRQ-RF |
| Acute and chronic respiratory failure with hypercapnia | J95.97 | AHRQ-RF |
| Respiratory disorders in diseases classified elsewhere | J95.98 | AHRQ-RF |
| Pulmonary insufficiency following trauma and surgery | J95.99 | AHRQ-RF |
| Acute postprocedural respiratory failure | J95.100 | AHRQ-RF |
| Acute respiratory failure with hypoxia | J95.101 | AHRQ-RF |
| Acute respiratory failure with hypercapnia | J95.102 | AHRQ-RF |
| Acute pulmonary insufficiency following thoracic surgery | J95.103 | AHRQ-RF |
| Acute pulmonary insufficiency following nonthoracic surgery | J95.104 | AHRQ-RF |
| Chronic pulmonary insufficiency following surgery | J95.105 | AHRQ-RF |
| Acute and chronic postprocedural respiratory failure | J95.106 | AHRQ-RF |
| Acute respiratory distress syndrome | J95.107 | AHRQ-RF |
| Chronic respiratory failure, unspecified whether with hypoxia or hypercapnia | J95.108 | AHRQ-RF |
| Chronic respiratory failure with hypoxia | J95.109 | AHRQ-RF |
| Chronic respiratory failure with hypercapnia | J95.110 | AHRQ-RF |
| Respiratory arrest | J95.111 | AHRQ-RF |
| Pneumonia due to streptococcus pneumonia | J95.112 | AHRQ-PNA |
| Other bacterial pneumonia | J95.113 | AHRQ-PNA |
| Pneumonia due to Pseudomonas | J95.114 | AHRQ-PNA |
| Pneumonia due to streptococcus | J95.115 | AHRQ-PNA |
| Pneumonia due to staphylococcus | J95.116 | AHRQ-PNA |
| Pneumonia due to Methicillin susceptible Staphylococcus | J95.117 | AHRQ-PNA |
| Pneumonia due to Methicillin susceptible Staphylococcus aureus | J95.118 | AHRQ-PNA |
| Pneumonia due to Escherichia coli | J95.119 | AHRQ-PNA |
| Pneumonia due to other Gram-negative bacteria | J95.120 | AHRQ-PNA |
| Pneumonia due to other specified bacteria | J95.121 | AHRQ-PNA |
| Unspecified bacterial pneumonia | J95.122 | AHRQ-PNA |
| Pneumonia due to other specified infectious organisms | J95.123 | AHRQ-PNA |
| Invasive pulmonary aspergillus | J95.124 | AHRQ-PNA |
| Bronchopneumonia, unspecified organism | J95.125 | AHRQ-PNA |
| Pneumonia, unspecified organism | J95.126 | AHRQ-PNA |
| Pneumonia due to Klebsiella pneumonia | J95.127 | AHRQ-PNA |
| Pneumonia due to other streptococci | J95.128 | AHRQ-PNA |
| Pneumonitis due to solids and liquids | J95.129 | AHRQ-ASP |
| Hypostatic pneumonia, unspecified organism | J95.130 | AHRQ-PNA |
| Other ill-defined and unknown causes of morbidity and mortality | J95.131 |  |
| Respiratory conditions due to chemical fumes and vapors | J95.132 |  |
| Pneumonitis due to solids and liquids | J95.133 | AHRQ-ASP |
| Pneumonitis due to inhalation of food and vomit | J95.134 | AHRQ-ASP |
| Postprocedural pneumothorax | J95.135 |  |
| Septic pulmonary embolism without acute cor pulmonale | J95.136 | AHRQ-PE |
| Other pulmonary embolism without acute cor pulmonale | J95.137 | AHRQ-PE |
| Air embolism following infusion, transfusion and therapeutic injection, initial encounter | J95.138 | AHRQ-PE |
| Complication of other artery following a procedure, not elsewhere classified, initial encounter | J95.139 |  |
| Complication of vein following a procedure, not elsewhere classified, initial encounter | J95.140 |  |
| Embolism due to cardiac prosthetic devices, implants and grafts, initial encounter | J95.141 | AHRQ-PE |
| Embolism due to cardiac prosthetic devices, implants and grafts, initial encounter | J95.142 | AHRQ-PE |
| Embolism due to vascular prosthetic devices, implants and grafts, initial encounter | J95.143 | AHRQ-PE |
| Embolism due to vascular prosthetic devices, implants and grafts, initial encounter | J95.144 | AHRQ-PE |

## References

1. Maron BA. Revised Definition of Pulmonary Hypertension and Approach to Management: A Clinical Primer. J Am Heart Assoc. 2023;12(8):e029024.

2. Ramakrishna G, Sprung J, Ravi BS, Chandrasekaran K, McGoon MD. Impact of pulmonary hypertension on the outcomes of noncardiac surgery: predictors of perioperative morbidity and mortality. J Am Coll Cardiol. 2005;45(10):1691–1699.

3. Price LC, Martinez G, Brame A, et al. Perioperative management of patients with pulmonary hypertension undergoing non-cardiothoracic, non-obstetric surgery: a systematic review and expert consensus statement. British journal of anaesthesia. 2021;126(4):774–790.

4. Kruthiventi SC, Kane GC, Sprung J, Weingarten TN, Warner ME. Postoperative pulmonary complications in contemporary cohort of patients with pulmonary hypertension. Bosn J Basic Med Sci. 2019;19(4):392–399.

5. Duchnowski P, Hryniewiecki T, Kusmierczyk M, Szymanski P. Right ventricular systolic pressure as a predictive factor for postoperative pneumonia in patients with valvular heart disease. Kardiol Pol. 2019;77(10):969–971.

6. Aasen DM, Bronsert MR, Rozeboom PD, et al. Relationships between predischarge and postdischarge infectious complications, length of stay, and unplanned readmissions in the ACS NSQIP database. Surgery. 2021;169(2):325–332.

7. Elixhauser A, Steiner C, Harris DR, Coffey RM. Comorbidity measures for use with administrative data. Med Care. 1998;36(1):8–27.

8. Protopapa KL, Simpson JC, Smith NC, Moonesinghe SR. Development and validation of the Surgical Outcome Risk Tool (SORT). Br J Surg. 2014;101(13):1774–1783.

9. Braunstein JB, Anderson GF, Gerstenblith G, et al. Noncardiac comorbidity increases preventable hospitalizations and mortality among Medicare beneficiaries with chronic heart failure. J Am Coll Cardiol. 2003;42(7):1226–1233.

10. Murad K, Kitzman DW. Frailty and multiple comorbidities in the elderly patient with heart failure: implications for management. Heart Fail Rev. 2012;17(4-5):581-588.

11. Thomas LE, Li F, Pencina MJ. Overlap Weighting: A Propensity Score Method That Mimics Attributes of a Randomized Clinical Trial. Jama. 2020;323(23):2417–2418.

12. Thomas L, Li F, Pencina M. Using Propensity Score Methods to Create Target Populations in Observational Clinical Research. Jama. 2020;323(5):466–467.

13. Hoeper MM, Humbert M, Souza R, et al. A global view of pulmonary hypertension. Lancet Respir Med. 2016;4(4):306–322.

14. Hill NS, Roberts KR, Preston IR. Postoperative Pulmonary Hypertension: Etiology and Treatment of a Dangerous Complication. Respiratory Care. 2009;54(7):958–968.

15. Margraf A, Ludwig N, Zarbock A, Rossaint J. Systemic Inflammatory Response Syndrome After Surgery: Mechanisms and Protection. Anesth Analg. 2020;131(6):1693–1707.

16. Koirala U, Thapa PB, Joshi MR, Singh DR, Sharma SK. Systemic Inflammatory Response Syndrome following Gastrointestinal Surgery. JNMA J Nepal Med Assoc. 2017;56(206):221–225.

17. Quarck R, Nawrot T, Meyns B, Delcroix M. C-reactive protein: a new predictor of adverse outcome in pulmonary arterial hypertension. J Am Coll Cardiol. 2009;53(14):1211–1218.

18. Baptista de Barros Ribeiro Dourado LP, Santos M, Moreira-Gonçalves D. Nets, pulmonary arterial hypertension, and thrombo-inflammation. J Mol Med (Berl). 2022;100(5):713–722.

19. Asllanaj B, Benge E, Bae J, McWhorter Y. Fluid management in septic patients with pulmonary hypertension, review of the literature. Front Cardiovasc Med. 2023;10:1096871.

20. Hansen L, Burks M, Kingman M, Stewart T. Volume Management in Pulmonary Arterial Hypertension Patients: An Expert Pulmonary Hypertension Clinician Perspective. Pulm Ther. 2018;4(1):13–27.

21. Ma G, Liao W, Qiu J, Su Q, Fang Y, Gu B. N-terminal prohormone B-type natriuretic peptide and weaning outcome in postoperative patients with pulmonary complications. The Journal of international medical research. 2013;41(5):1612–1621.

22. Rosenkranz S, Pausch C, Coghlan JG, et al. Risk stratification and response to therapy in patients with pulmonary arterial hypertension and comorbidities: A COMPERA analysis. J Heart Lung Transplant. 2023;42(1):102–114.

23. Reesink HJ, Tulevski, II, Marcus JT, et al. Brain natriuretic peptide as noninvasive marker of the severity of right ventricular dysfunction in chronic thromboembolic pulmonary hypertension. The Annals of thoracic surgery. 2007;84(2):537–543.

24. Yap LB, Ashrafian H, Mukerjee D, Coghlan JG, Timms PM. The natriuretic peptides and their role in disorders of right heart dysfunction and pulmonary hypertension. Clin Biochem. 2004;37(10):847–856.

25. Messmer AS, Zingg C, Müller M, Gerber JL, Schefold JC, Pfortmueller CA. Fluid Overload and Mortality in Adult Critical Care Patients-A Systematic Review and Meta-Analysis of Observational Studies. Critical care medicine. 2020;48(12):1862–1870.

26. Price LC, Montani D, Jais X, et al. Noncardiothoracic nonobstetric surgery in mild-to-moderate pulmonary hypertension. Eur Respir J. 2010;35(6):1294–1302.

27. Minai OA, Venkateshiah SB, Arroliga AC. Surgical intervention in patients with moderate to severe pulmonary arterial hypertension. Conn Med. 2006;70(4):239–243.

28. Canet J, Gallart L, Gomar C, et al. Prediction of postoperative pulmonary complications in a population-based surgical cohort. Anesthesiology. 2010;113(6):1338–1350.

29. Lawrence VA, Dhanda R, Hilsenbeck SG, Page CP. Risk of pulmonary complications after elective abdominal surgery. Chest. 1996;110(3):744–750.

30. Maron BA, Brittain EL, Hess E, et al. Pulmonary vascular resistance and clinical outcomes in patients with pulmonary hypertension: a retrospective cohort study. Lancet Respir Med. 2020;8(9):873–884.

31. Chung L, Liu J, Parsons L, et al. Characterization of connective tissue disease-associated pulmonary arterial hypertension from REVEAL: identifying systemic sclerosis as a unique phenotype. Chest. 2010;138(6):1383–1394.

32. Humbert M, Kovacs G, Hoeper MM, et al. 2022 ESC/ERS Guidelines for the diagnosis and treatment of pulmonary hypertension. European heart journal. 2022;43(38):3618–3731.

